# Physical determinants of daily physical activity in older men and women

**DOI:** 10.1101/2024.04.15.24305391

**Authors:** Laura Karavirta, Timo Aittokoski, Katja Pynnönen, Timo Rantalainen, Kate Westgate, Tomas Gonzales, Lotta Palmberg, Joona Neuvonen, Jukka A. Lipponen, Katri Turunen, Riku Nikander, Erja Portegijs, Taina Rantanen, Søren Brage

## Abstract

**Introduction:** The ability to perform bodily movement varies in ageing men and women. We investigated whether physical fitness may explain sex differences in daily physical activity energy expenditure (PAEE) among older people.

**Methods:** A population-based cohort of 75, 80, and 85-year-old men and women (n=409, 62 % women) underwent laboratory-based assessment of walking speed, maximal knee extension strength, and body fat percentage. Free-living physical activity was assessed as total PAEE, and light (LPA) and moderate-to-vigorous physical activity (MVPA) using individually calibrated combined accelerometry and heart rate sensing. Path modelling was used to examine indirect associations between sex, physical fitness, and physical activity.

**Results:** Men had a more favourable physical fitness profile and higher overall PAEE (mean 34.0 (SD 10.8) kJ/kg/day) than women (28.3 (8.4) kJ/kg/day, p<0.001). The path model for PAEE explained 33 % of the variance. The direct association of sex on PAEE was non-significant, whereas the association between sex and PAEE through body fat (β=0.20, p<0.001) and walking speed (β=0.05, p=0.001) were statistically significant. Similarly, the associations between sex and MVPA through body fat (β=0.11, p=0.002) and walking speed (β=0.05, p=0.001) were significant, as were the associations between sex and LPA through body fat (β=0.24, p<0.001) and walking speed (β=0.03, p=0.019).

**Conclusion:** Differences in physical activity between men and women may reflect underlying differences in cardiorespiratory fitness and adiposity. These results highlight the importance of maintaining physical fitness to support active living in older adults.

## Introduction

Low physical activity is of concern, especially in older adults, who are at risk for functional limitations and loss of independence (Westerterp, 2000). The overall volume of physical activity is typically expressed as daily physical activity energy expenditure (PAEE), which is accumulated through the engagement of behaviours of different intensities. Wearable sensors, such as accelerometers and heart rate (HR) sensors, can be used to objectively assess physical activity and estimate PAEE (Brage et al., 2015; Ceesay et al., 1989). To convert data from wearables into intensity estimates, often grouped into light, moderate, and vigorous physical activity, methodological studies with suitable criterion measures and population groups have been conducted, validating both intensity and volume estimates of activity (Brage et al., 2004; Westgate et al., 2024). The implementation of wearables and the use of an inferential framework for PAEE estimation allows the evaluation of the joint health associations for activity volume and intensity as recently demonstrated for mortality in the UK Biobank (Strain et al., 2020). These prospective studies provide evidence of the health consequences of different activity behaviour profiles but say little about the determinants of physical activity, including individual physical characteristics, which decline at an accelerated rate in older people (Fleg et al., 2005).

Physical fitness is a group of physical characteristics that are defined as the “physiologic attribute determining a person’s ability to perform muscle-powered work”, whereas physical activity is bodily movement produced by skeletal muscles that results in energy expenditure (2018 Physical Activity Guidelines Advisory Committee, 2018). Therefore, instantaneous PAEE, or intensity, is dependent on the individual capacity to perform muscle-powered work at any given moment. Physical fitness is a multicomponent characteristic including body composition, cardiorespiratory fitness, and muscle strength, which are all independently associated with health (Blair et al., 1996; Jayanama et al., 2022; Li et al., 2018). It is well known that men have a more favourable body composition, and higher muscle strength and cardiorespiratory fitness compared to women. The mean difference in cardiorespiratory fitness between men and women is approximately 1 to 2 METs in older age (Fleg et al., 2005; Ogawa et al., 1992), which could translate to higher physical activity intensity and volume in men compared to women. Thus, the previously observed tendency of men to favour vigorous intensity activities more than women, and women to engage in lower intensity activities more often compared to men (Lindsay et al., 2019; Sattelmair et al., 2011)may be by constraint rather than choice.

The aim of the study was to explore the direct and indirect associations between physical characteristics and physical activity volume and intensity among older people. More specifically, we examined how the components of physical fitness (body fat percentage, walking speed, and muscle strength) may mediate the relationships between sex and physical activity. We hypothesized that some of the variance in physical activity is explained by physical fitness rather than sex.

## Methods

### Participants

The data for the present analyses are from a population-based observational study including three age cohorts: 75, 80, and 85 years and we have published the study protocol previously (Rantanen et al., 2018). The personal details such as sex and date of birth, were available in the sample specifics drawn from the Digital and Population Services Agency (https://dvv.fi/en). We targeted everyone living independently near the city centre of Jyväskylä, in Central Finland, who were born during the years specified. The ethical committee of the Central Finland Health Care District provided a statement on the AGNES study protocol on the 23^rd^ of August, 2017. Participants were required to provide a written informed consent.

Of the 2791 people approached, the overall participation rate was 36.6 (Portegijs et al., 2019). From this sample of 1021 participants (57.3 % women), 910 individuals agreed to participate in the laboratory assessments and were thus also invited to take part in the device-based physical activity monitoring in free living (Fig. 1). To those who agreed to wear a thigh-mounted accelerometer (n=495), we additionally offered an electrocardiogram (ECG) recorder unless the participant had an active implantable medical device such as a heart pacemaker (n=19). We excluded participants with less than 3 days of wearables data and were able to obtain combined accelerometry and HR data for 409 participants (61.9 % women).

**Fig. 1.**
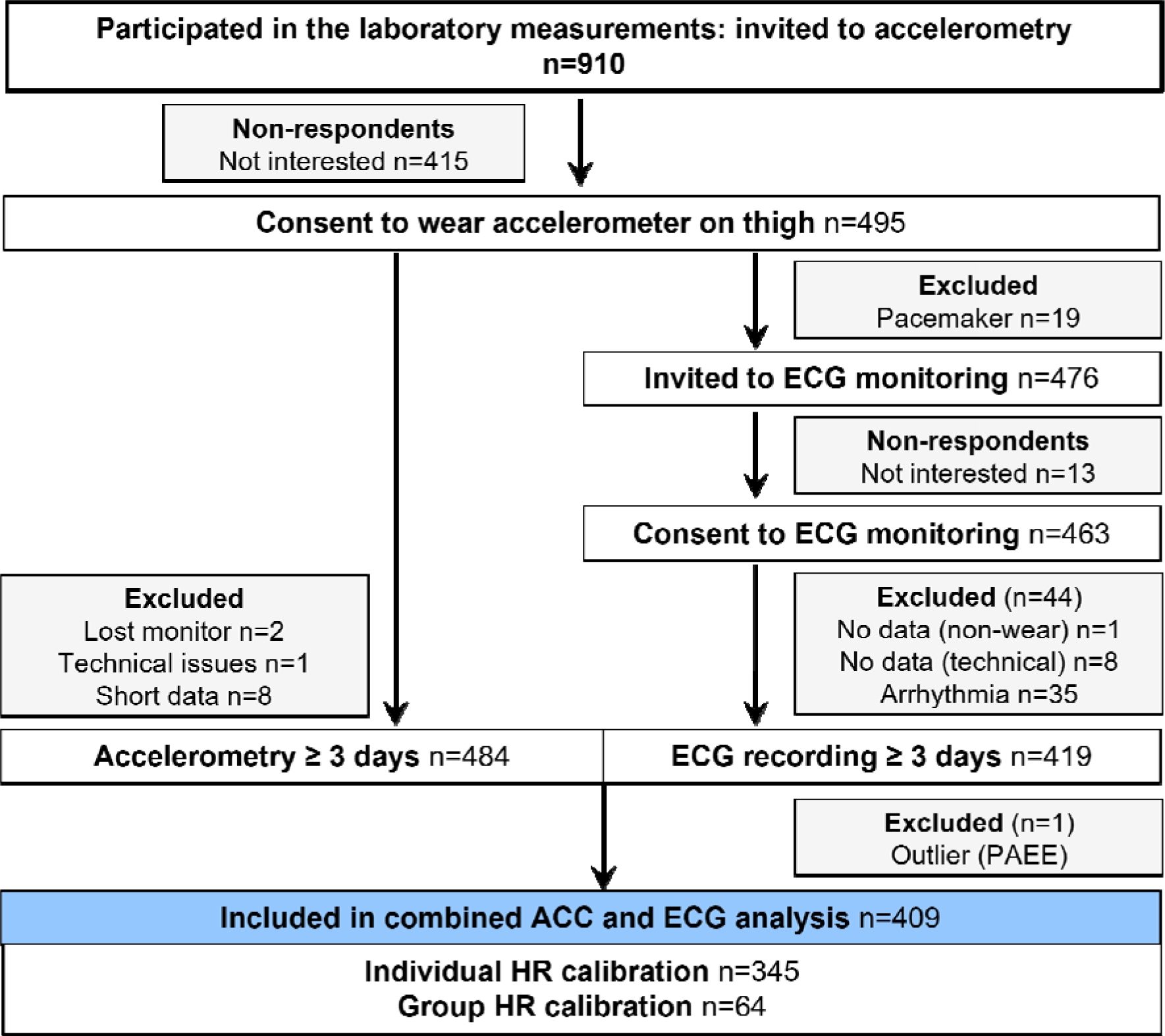
Participant flow chart. Combined wearable sensor data from the AGNES study at baseline. ACC, accelerometry; ECG, electrocardiogram; PAEE, physical activity energy expenditure; HR, heart rate.

### Physical fitness measurements

#### Six-minute walk test (6MWT)

We used the 6-minute walk test to assess cardiorespiratory fitness and to individually calibrate HR to PAEE. The walking test was performed at the research centre on an indoor 20-meter corridor at a self-selected usual pace (Karavirta et al., 2020). Walking speed (in km/h) was calculated from the total distance walked in six minutes. The participants wore the same sensors as during the free-living monitoring: a triaxial accelerometer (sampling continuously at 100 Hz, 13-bit, ±16 g, UKK RM42; UKK Terveyspalvelut Oy, Tampere, Finland) and an ECG recorder (14-bit, ±16 g, 250 Hz, eMotion Faros 180, Bittium Corporation, Oulu, Finland). HR was derived from the ECG recordings using a built-in QRS detection of Kubios HRV Premium 3.2.0 software (Kubios Oy, Kuopio, Finland) that is based on the Pan-Tompkins algorithm (Pan & Tompkins, 1985). Any artifacts were corrected using automatic noise detection with manual editing, when necessary. Walking HR was calculated by averaging the resulting RR intervals (the time interval between two successive R waves on the ECG) over the last minute of the test and converting to HR in beats per minute (bpm). Recovery HR after the test was analysed from the RR intervals over 90 seconds, subjected to quadratic regression against recovery time, and solved for 45 seconds, similar to previous work (Brage et al., 2007; Westgate et al., 2024). For the PAEE estimation, HR values were expressed above sleeping HR (HRaS) but are reported as absolute values in the results for readability.

#### Maximal knee extension strength

Knee extension strength was measured at an angle of 60 degrees from the fully extended leg towards flexion. Following a practice trial, the test was performed at least three times allowing one minute rest between trials until no further improvement occurred (Rantanen et al., 2018). The highest value in newtons normalized to body mass to account for the strength requirements of transferring the body (Davies & Dalsky, 1997) was recorded as the test result. The test-retest reliability of the test is excellent; for measurements in 80-year-olds performed 1–2 weeks apart, the Pearson correlation coefficient was 0.965 (Rantanen et al., 1997).

#### Adiposity

Multi-frequency bioelectrical impedance measurement (InBody 720, Biospace, Seoul, Korea) was used to assess adiposity as body fat percentage. Measurements were performed with participants wearing light clothing standing barefoot on the device platform and holding the handles in both hands. Physical assessments in the laboratory also included standard objective anthropometric measurements of height and body mass.

### Physical activity monitoring during free-living

Physical activity monitoring took place between a home interview and a laboratory visit, at which the wearable sensors were attached and removed, respectively. The thigh-worn triaxial accelerometer was taped on by a research assistant to the anterior aspect of the mid-thigh of the dominant leg, and the ECG recorder was attached with an adhesive strip that included two electrodes 12 centimetres apart. The strip was attached either on the sternum or diagonally on the left side of the chest under the breast to ensure comfortable wear depending on the anatomy of the participant. Both monitors were covered with a self-adhesive film for waterproofing to enable constant wear including during showering. However, swimming or bathing was discouraged during the monitoring period. The electrode and adhesives were replaced once in the middle of the monitoring period by a research assistant.

#### Accelerometry and ECG signal processing

Raw acceleration data were calibrated to local gravity, based on the principle described elsewhere (van Hees et al., 2014). However, we deviated slightly from this procedure by including only gain and offset in the optimization procedure and utilizing the Levenberg-Marquardt algorithm in the iterative minimization of the error function (Karavirta et al., 2020). From resulting gravity-calibrated acceleration values, mean amplitude deviation (MAD = 1/n *∑ |r*_k_* – r|) was calculated from the vector magnitude (Euclidian norm) of the resultant acceleration (√X^2^+Y^2^+Z^2^) in nonoverlapping 5-second epochs (Vähä-Ypyä et al., 2015). This activity-related acceleration metric is robust to residual calibration error through the subtraction operation. MAD was then resampled to 10-second epochs to match the 10-second averaging of the HR time series. The data were visually checked day-by-day to ensure that only days with complete 24-hour data without non-wear were included in the subsequent analysis. Data of 11 participants owing to either loss of monitor (n□=□2), technical error (n□=□1), or data availability for less than three full days (n□=□8) were excluded (Fig.1).

Free-living ECG recordings were analysed with commercial medically certified Awario arrhythmia analysis algorithms (Awario, Heart2Save, Kuopio, Finland) (Santala et al., 2022). The algorithm detects and removes noisy segments of the ECG, detects the QRS complexes, and estimates the average HR in a 10-second sliding time window from the analysable parts of the ECG. Participants with persistent atrial fibrillation throughout the recording were excluded from further analyses (n=35, Fig. 1). Additionally, one outlier was excluded due to unreliable HR resulting in a PAEE estimate more than 3 SD above the sample mean.

#### Estimation of physical activity energy expenditure

PAEE was estimated from the 10-second epoch time series of MAD and HR separately, and then combined in a branched equation model (Brage et al., 2004). A linear regression equation for estimating instantaneous PAEE (intensity) from MAD was established using treadmill walking in a separate dataset of 12 older participants (methods described in Supplement 1):

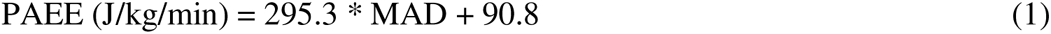

The linear equation was used for all accelerometry epochs where MAD was above a flex movement point (Brage et al., 2007). Flex acceleration was defined as the group mean acceleration during the slowest speed of the treadmill walking (1.5 km/h) which is almost equal to the previously used flex point defined as 50 % of the acceleration measured during walking at 3.2 km/h (Brage et al., 2007). Between the flex point and zero acceleration (the lowest recorded MAD during free living), PAEE was extrapolated linearly between PAEE at the flex point and the origin of 0 g, 0 J/kg/min (Supplement 1, Fig.S1.1).

From the HR time-series data, PAEE was estimated using an individual calibration equation. Individual variation in the relationship between HR and energy expenditure was captured using a novel calibration method based on a self-paced walking test (Westgate et al., 2024). The method can be used to individually determine the linear relationship between HR and PAEE, which can then be used to compute PAEE at any HR level as follows:

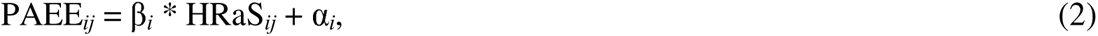

where PAEE*_ij_* and HRaS*_ij_* are physical activity energy expenditure (J/kg/min) and heart rate (bpm above sleeping heart rate) for participant *i* for epoch *j*. β*_i_* and α*_i_* are the slope and intercept of the linear regression equation for participant *i*.

For calculating the individual slope and intercept of the linear equation, we computed one more parameter from the 6-minute walk test in addition to walking and recovery HR: energy pulse, which was defined as the average HR above sleep divided by the energy cost of walking using a previously validated equation (Ludlow & Weyand, 2016):

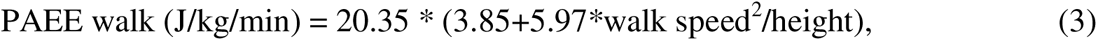

where 20.35 is the caloric equivalent of oxygen in J/ml O_2_ (Consolazio et al., 1963), walk speed is in m/s, and height is in m. Energy pulse was further transformed using a natural logarithm (ln EP). We used the combination of the simple and complex equations by Westgate and colleagues (Westgate et al., 2024) with an average weighting (factor 0.5) to account for beta-blocker use and sex but to avoid parameter overfitting:

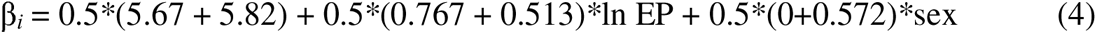

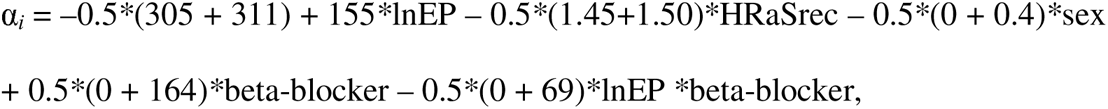

where *i* indicates participant *i*, sex was coded 0=woman and 1=man, and beta-blocker use 0=no and 1=yes. HRaSrec is recovery heart rate 45 seconds after the walking test above sleeping HR.

The individual slope β*_i_* and intercept α*_i_* were used to estimate PAEE when HR was at least 5 bpm above flex HR. Flex HR is a theoretical deflection point above which HR is linearly associated with PAEE and is classically defined as “the mean of the highest HR during rest and the lowest HR during the lightest imposed exercise” (Ceesay et al., 1989). Below this point, PAEE was interpolated to 0 J/kg/min at resting HR, since at low HR levels, HR can fluctuate due to other factors than physical activity (Brage et al., 2007; Westgate et al., 2024). Flex HR was individually estimated from the midpoint between the HR corresponding to PAEE at the slowest walking speed of our treadmill protocol (108 J/kg/min at 1.5 km/h, Supplement 1) and resting HR determined as 10 bpm above sleeping HR. For participants in whom we were unable to extract valid HR from the walking test (n=64, Fig.1), we used a group calibration equation based on all participants in the study with valid calibration whilst accounting for age, sex, beta blockage, and sleeping HR:

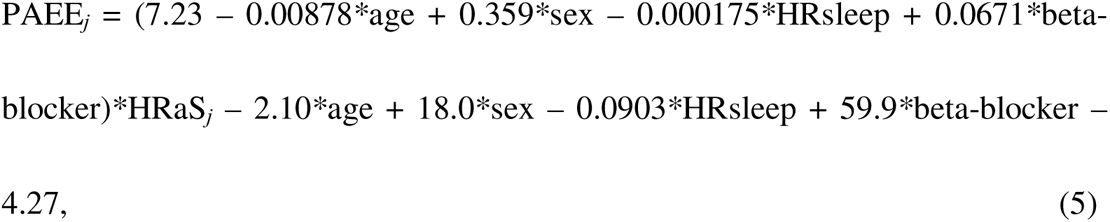

where HRaS*_j_* is HR in epoch *j* expressed as above sleeping HR. In the group-calibrated subsample, flex HR (above sleep) was predicted from sleeping HR: 0.37 * sleeping HR - 2.5 + 5 (Brage et al., 2007). Sleeping HR was defined as the median of the 10^th^ lowest HR observed during sleep across multiple 24-hour periods (Brage et al., 2004).

Accelerometry and HR-based PAEE time series were combined using a branched equation modelling (Brage et al., 2004). In essence, the model gives a larger weight for accelerometry-based PAEE estimation when acceleration and HR are low (due to the known fluctuation in HR at low levels regardless of physical activity), and a larger weight for HR when it is above the flex point and accompanied by physical movement, implemented as described elsewhere (Brage et al., 2015). In addition, whenever HR was not available due to noise or transient arrhythmia in the ECG signal, acceleration-PAEE was used. HR availability is reported as a percentage of the accelerometer wear time.

In addition to overall PAEE, we calculated PAEE at moderate-to-vigorous (MVPA, 2–5 net METs) and light intensity (LPA, 0.5–2 net METs) as minutes and proportions of the overall PAEE. MVPA was also expressed in MET-minutes. The distribution of PAEE was additionally calculated in fine-grained intensity bins of 0.5 to 1.0 METs each. One MET was defined according to the standard of 3.5 ml/kg/min (Ainsworth et al., 2011) and converted to J/kg/min using 20.35 J/ml (Consolazio et al., 1963), resulting in a value of 71 J/min/kg.

### Covariates

Age, duration of education, chronic conditions, beta-blocker use, smoking, accelerometer wear time, and HR availability as a percentage of the accelerometer wear time were tested as potential covariates. The duration of education (years) and the total number of self-reported physician-diagnosed chronic conditions were self-reported during the home interview (Rantanen et al., 2018). Participants reported their current medication in a postal questionnaire, and beta-blocker medication was identified (yes/no) based on the ATC (Anatomical Therapeutic Chemical) code. Smoking was self-reported as never, past, or current, and dichotomously reclassified as never or past/present.

### Statistical analyses

Group values are means followed by standard deviations (SD). The differences in participant characteristics and physical activity between the sexes were tested using an independent-samples t-test and Cohen’s d for effect sizes. Cross-tabulation and chi-squared tests were used for categorical variables. Bivariate associations among components of physical fitness and physical activity, stratified by sex, were analysed using Pearson’s correlation. Path modelling was conducted to examine whether the components of physical fitness (body fat, walking speed, and muscle strength) mediate the relationships between sex and physical activity. The model is structured considering the previously observed heterogeneity of physical fitness within sexes which suggests that stratification by sex alone may not account for the individual variance in physical activity. Maximum likelihood robust (MLR) estimator was used to obtain parameter estimates supposing missing values to be missing at random (MAR). The covariance coverage was 0.968 at minimum. In separate models, a component of physical activity (i.e., overall PAEE, MVPA, or LPA) was an outcome and sex was an explanatory variable. All three physical fitness components were added simultaneously to the models to test the potential mediator effect, and they were allowed to correlate with each other. Each component of physical fitness and physical activity was adjusted for age, duration of education, and number of chronic conditions. Additionally (or based on modification indexes), age was allowed to correlate with the number of chronic conditions and the duration of education. In the final models, only statistically significant associations were included. Chi-square (χ2), root mean squared error of approximation (RMSEA, <0.06), comparative fit index (CFI, >0.95), Tucker-Lewis Index (TLI, >0.95), standardized root mean square residual (SRMR <0.08) are reported as indices of model fit.

As a sensitivity analysis and to enhance the applicability of the present results to studies that use an accelerometer only, path modelling was also performed for accelerometry-based PAEE without HR sensing. Statistical significance was set at p<0.05. Analysis was conducted using IBM SPSS Statistics 28.0.1.1 and Mplus version 8.6 (Muthén & Muthén, 2017).

## Results

Men and women in the present sample did not significantly differ in terms of age, the number of chronic conditions, duration of education, or beta-blocker use (table 1). As expected, men had larger body size with lower body fat percentage compared to women. Muscle strength and walking speed were higher in men. Women had higher HR during sleep, self-paced walking, and recovery after the walk compared to men.

**Table 1.** AGNES study participant characteristics.

|  | Women<br>(n=253) |  | Men<br>(n=156) |  | Total<br>(n=409) |  |  |
| --- | --- | --- | --- | --- | --- | --- | --- |
|  | Mean | SD | Mean | SD | Mean | SD | p (sex) |
| Age | 78.2 | 3.4 | 78.2 | 3.2 | 78.2 | 3.3 | 0.985 |
| Body height (m) | 1.58 | 0.05 | 1.72 | 0.06 | 1.64 | 0.09 | <0.001 |
| Body mass (kg) | 70.1 | 11.8 | 79.1 | 11.3 | 73.5 | 12.4 | <0.001 |
| Fat Free Mass (kg) | 42.1 | 4.5 | 57.2 | 6.9 | 47.9 | 9.2 | <0.001 |
| Body Fat (%) | 39.0 | 7.1 | 27.0 | 6.7 | 34.5 | 9.1 | <0.001 |
| Chronic conditions (count) | 3.0 | 1.8 | 2.7 | 1.8 | 2.9 | 1.8 | 0.17 |
| Education (years) | 11.6 | 4.0 | 12.1 | 4.4 | 11.8 | 4.2 | 0.31 |
| Muscle strength (N/kg) | 4.23 | 1.23 | 5.70 | 1.45 | 4.78 | 1.50 | <0.001 |
| Walk Speed (km/h) | 4.13 | 0.79 | 4.45 | 0.76 | 4.25 | 0.79 | <0.001 |
| Walk EE estim. (net stdMET) | 2.57 | 0.52 | 2.65 | 0.50 | 2.60 | 0.51 | 0.10 |
| HR walk (bpm) | 104.9 | 15.3 | 98.6 | 16.0 | 102.5 | 15.8 | <0.001 |
| HR rec (bpm) | 88.8 | 15.0 | 84.4 | 14.6 | 87.1 | 15.0 | 0.007 |
| HR sleep (bpm) | 52.5 | 6.7 | 48.9 | 6.5 | 51.2 | 6.8 | <0.001 |
|  | n | (%) | n | (%) | n | (%) |  |
| Past or present smoking (yes) | 35 | 13.8 | 67 | 43.2 | 102 | 25.0 | <0.001 |
| Betablocker use (yes) | 91 | 36.0 | 45 | 28.8 | 136 | 33.3 | 0.138 |

EE estim., estimated energy expenditure; HR, heart rate; rec; recovery

Figure 2 presents overall PAEE stratified by age and sex, showing inverse associations for age in both sexes. The allocation of PAEE to LPA and MVPA as energy expenditure, proportion, and minutes per day are presented in Table 2. Approximately half of the daily PAEE was spent in light-intensity activities in both men and women (p=0.44), while the contribution of activities of at least moderate intensity was 28% in men and 23% in women (p<0.001). All the other indicators of daily physical activity were also higher in men compared to women (p<0.001). The effect size of sex on PAEE varied across the intensity spectrum ranging from 0.25 to 0.56, being largest at 1.5-2.0 METs (Fig.3). PAEE, MVPA, and LPA were significantly associated with body fat percentage, walking speed, and muscular strength in both men and women (Table 3).

**Fig 2.**
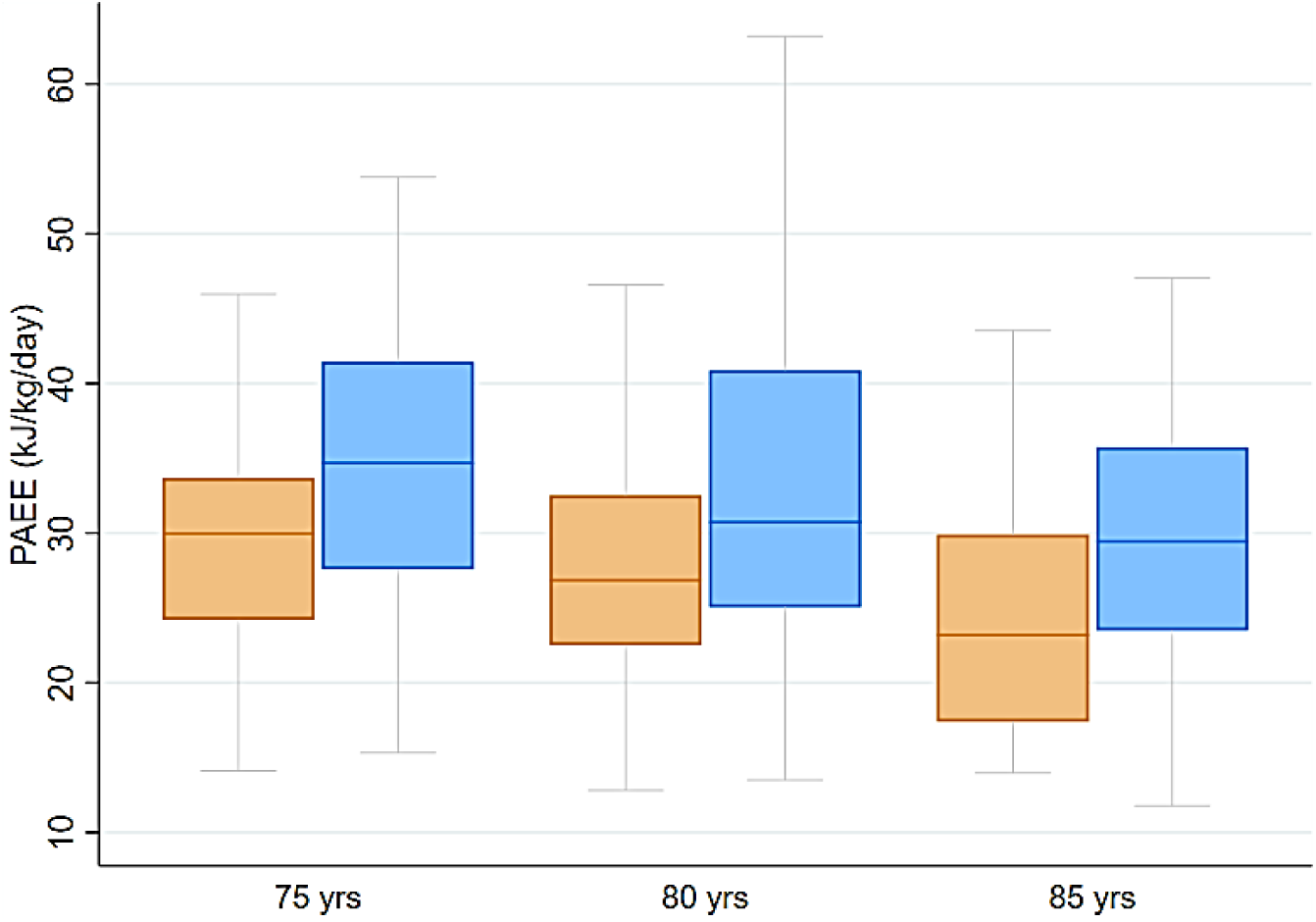
A boxplot of the median (±first and third quartile with lower and upper adjacent values) of daily physical activity energy expenditure stratified by age group and sex (women in orange, men in blue).

**Fig 3.**
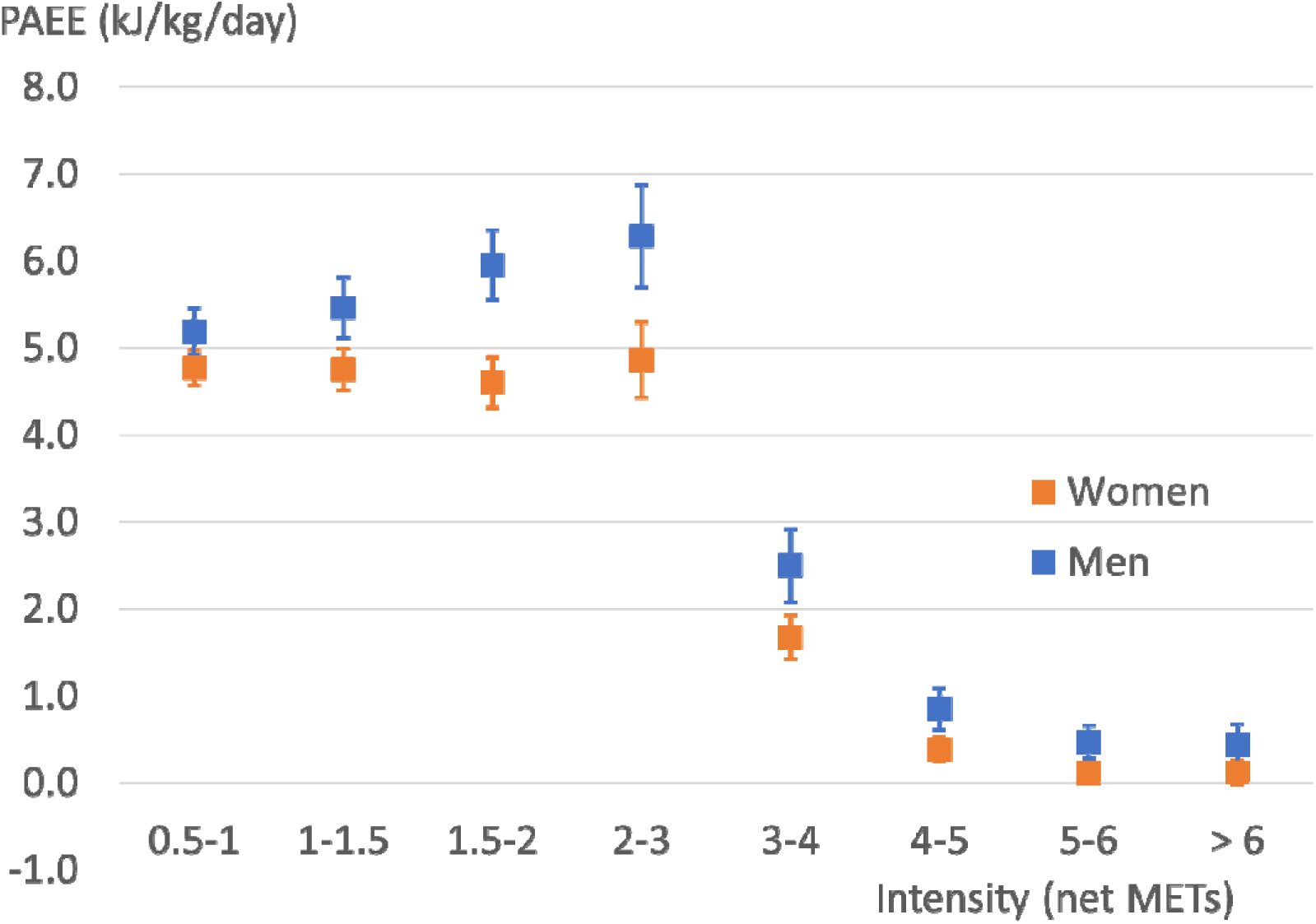
Daily accumulation of physical activity energy expenditure (PAEE, mean ± 95 % CI) in men and women at different intensity levels expressed as standard metabolic equivalents above the resting level (net METs). Note, bin size differences below and above 2 net MET.

**Table 2.** Physical activity metrics stratified by sex.

|  | Women |  | Men |  | Total |  |  |
| --- | --- | --- | --- | --- | --- | --- | --- |
|  | Mean | SD | Mean | SD | Mean | SD | p (sex) |
| PAEE (kJ/kg/day) | 28.3 | 8.4 | 34.0 | 10.8 | 30.5 | 9.8 | <0.001 |
| PAEE from MVPA (kJ/kg/day) | 7.1 | 5.7 | 10.5 | 7.5 | 8.4 | 6.6 | <0.001 |
| PAEE from MVPA (%) | 22.7 | 12.5 | 28.0 | 13.1 | 24.7 | 13.0 | <0.001 |
| PAEE from LPA (kJ/kg/day) | 14.1 | 4.8 | 16.6 | 5.4 | 15.1 | 5.2 | <0.001 |
| PAEE from LPA (%) | 50.0 | 8.7 | 49.3 | 8.5 | 49.7 | 8.6 | 0.44 |
| LPA (min/day) | 184 | 59 | 211 | 66 | 194 | 63 | <0.001 |
| MVPA (min/day) | 37 | 27 | 52 | 32 | 43 | 30 | <0.001 |
| MVPA (min/week) | 260 | 187 | 364 | 227 | 299 | 209 | <0.001 |
| MVPA (net stdMET mins/week) | 703 | 556 | 1035 | 741 | 829 | 652 | <0.001 |
| Wear time (days) | 7.2 | 1.1 | 7.1 | 1.2 | 7.1 | 1.1 | 0.56 |
| HR availability (%) | 74.8 | 24.4 | 74.1 | 23.8 | 74.5 | 24.1 | 0.78 |

PAEE, physical activity energy expenditure; MVPA, moderate-to-vigorous physical activity; LPA, light physical activity; stdMET, standard metabolic equivalent (3.5 ml/kg/min)

**Table 3.** Pearson’s correlation coefficients between physical fitness and activity stratified by sex.

|  |  | PAEE<br>(kJ/kg/day) | PAEE from MVPA<br>(kJ/kg/day) | PAEE from LPA<br>(kJ/kg/day) |
| --- | --- | --- | --- | --- |
| *Women* |  |  |  |  |
| Body Fat (%) | r | **-0.429** | **-0.323** | **-0.381** |
|  | p | <0.001 | <0.001 | <0.001 |
| Walk speed (km/h) | r | **0.445** | **0.425** | **0.282** |
|  | p | <0.001 | <0.001 | <0.001 |
| Muscle strength (N/kg) | r | **0.401** | **0.401** | **0.224** |
|  | p | <0.001 | <0.001 | <0.001 |
| *Men* |  |  |  |  |
| Body Fat (%) | r | **-0.414** | **-0.341** | **-0.371** |
|  | p | <0.001 | <0.001 | <0.001 |
| Walk speed (km/h) | r | **0.454** | **0.440** | **0.302** |
|  | p | <0.001 | <0.001 | <0.001 |
| Muscle strength (N/kg) | r | **0.275** | **0.253** | **0.187** |
|  | p | 0.001 | 0.002 | 0.021 |

Sex had a bivariate association with overall PAEE (β=0.28; 95% CI 0.19-0.37; p<0.001), LPA (0.23; 0.14-0.33; p<0.001), and MVPA (0.25; 0.15-0.34; p<0.001). Results of the path model further indicated that men had a lower percentage of body fat than women but faster walking speed and higher maximal strength (Fig.4). All associations of the path model are presented in Supplementary file 2, Table S2.1. Of the physical fitness components, higher body fat percentage was negatively and higher walking speed positively associated with PAEE and MVPA while the associations of maximal strength with PAEE at different intensities were not significant. The direct effect of sex on PAEE at different intensities attenuated to non-significant.

**Fig 4.**
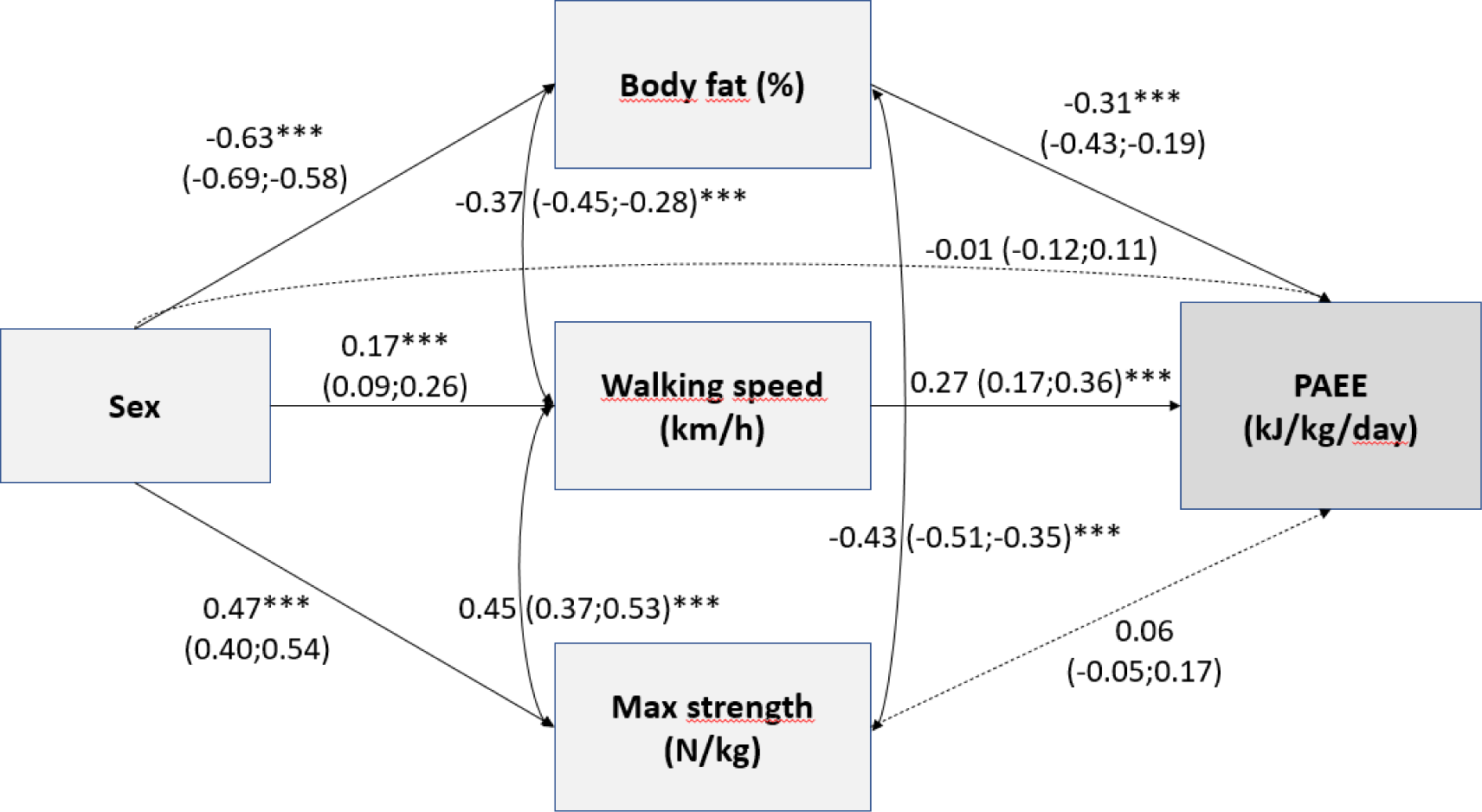
Standardized beta coefficients (with 95% confidence intervals) from path model testing factors describing physical fitness (body fat percentage, walking speed, and muscle strength) as mediators in the relationships between sex and physical activity energy expenditure (PAEE, *p≤0.05, **p≤0.01, ***p≤0.001).

The indirect effects from sex to PAEE through body fat (β=0.20, p<0.001) and walking speed (β=0.05, p=0.001) were statistically significant. The model explained 33.3 % of the variance in PAEE, and the model fit was good (χ2(9)=17.147, p=0.047, RMSEA 0.047, CFI 0.989, TLI 0.974, SRMR 0.047). Similarly, the indirect effects from sex to MVPA (Supplementary file 3 Fig.S3.1; Table S2.1) and from sex to LPA (Fig.S3.2; Table S2.1) through body fat and walking speed were statistically significant, and the models explained 27.2 % of the variance in MVPA and 21.1 % of the variance in LPA. Considering covariates, the number of chronic conditions was negatively related to all physical activity variables, walking speed, and muscle strength but positively associated with body fat. Duration of education was positively associated with walking speed and negatively with age, and the number of chronic conditions was positively associated with age.

In the sensitivity analysis where only accelerometry was used (Table S2.2; Fig.S3.3), all indirect effects from sex to accelerometry-based PAEE through the physical fitness components were significant including muscle strength: body fat (β=0.16, p<0.001), walking speed (β=0.05, p=0.002), and muscle strength (β=0.05, p=0.042) explaining 30.6 % of the variance in accelerometry-based PAEE (χ2(9)=16.075, p=.065, RMSEA 0.044, CFI 0.990, TLI 0.976, SRMR 0.045).

## Discussion

In this study, we investigated the physical determinants of PAEE in older Finnish men and women using combined accelerometry and heart rate sensing. The PAEE component from accelerometry was based on incremental walking data of a similar age group, and the PAEE component from heart rate was individually calibrated using a novel method based on self-paced submaximal walking. This is the first time the self-paced walking calibration was applied to older people, for whom it is a feasible method due to their heterogeneous functional ability. We observed approximately 20 % higher PAEE among older men compared to women, and our path analysis suggests this difference is largely explained by men’s lower body fat percentage and higher walking speed. Body fat percentage was an important contributor to LPA, whereas walking speed was the most important fitness component of MVPA. The results illustrate the intertwined association between physical fitness and physical activity in the context of ageing.

Previous studies have reported similar differences between younger men and women. In the Fenland study including the age range of 29 – 64 years, higher overall PAEE was observed in men (59 (SD 23) kJ/kg/day) compared to women (50 (20) kJ/kg/day) (Lindsay et al., 2019). Schrack et al. quantified physical activity both in absolute and relative terms and concluded, that based on the absolute activity counts women appeared less active, whereas in relative terms (percentage of HR reserve) women were less sedentary and engaged in more light and moderate activity compared to men (Schrack et al., 2018). They also observed that walking speed over 400 m was lower in the tertile with the lowest agreement between absolute and relative physical activity, which indirectly suggests that slow walking speed may be a determinant of the lower absolute volume of activity of a person who engages in activities at their own comfortable pace. Thus, the chosen methodology affects the observed difference between the sexes, even though the contribution of the distinct methodology for relative and absolute intensity quantification may also play a role (Schrack et al., 2018).

The majority of physical activity of older people is composed of walking, either incidental or planned. Therefore, the intensity at which people walk in their daily lives may have a major impact on the accumulation of MVPA. It can be inferred from the present and previous studies, that the preferred intensity of walking may be at either side of the common MVPA cut point of 3 METs (i.e. 2 net METs) (Rejeski et al., 2016; Schrack et al., 2012). In the present sample of older adults, the average walking intensity in the laboratory test was 2.6 (SD 0.5) net METs varying from 1.2 to 4.2 METs. Comparable walking intensity has also been reported previously when oxygen uptake was measured at “usual comfortable pace” in a diverse age group of 30–100 years. They walked on average at 13.0 (SD 2.8) ml/kg/min, which equals 2.7 standard net METs (Schrack et al., 2012). Even though the preferred speed may be higher in the laboratory than in a free-living environment (Takayanagi et al., 2019), those with lower walking speed in the laboratory are probably less likely to exceed the moderate intensity cut point while walking in free-living. For LPA, an even more important contributor was body fat percentage. This may be explained by the differential metabolic contributions of muscle and fat tissue on energy expenditure during physical activity. PAEE is generally expressed relative to total body mass, as was the case also in this study, which means that individuals with higher body fat percentage have less muscle mass for generating bodily movement, and more fat mass to carry during weight-bearing activities. The contribution of LPA was large, approximately half of the total PAEE, which may explain the importance of body composition for LPA.

We used combined sensing of accelerometry and HR for PAEE assessment to take advantage of both methods: 1) the well-established linear relationship between HR and energy expenditure across activity modes at moderate-to-vigorous intensities (Ceesay et al., 1989), and 2) the capability of thigh-mounted accelerometry to separate movement from non-movement at low intensities (Vähä-Ypyä et al., 2015), where the correlation between HR and PAEE is low (Ceesay et al., 1989). It has previously been shown that locomotion-based accelerometry models of PAEE may underestimate PAEE in free-living whereas individually calibrated HR models agree with doubly-labelled water measures of PAEE on average with high individual variance, and combined estimates being unbiased and with lower error variation (Brage et al., 2015). That said, correlations between combined sensing estimates and accelerometry-only estimates are relatively high, and our sensitivity analysis showed that the present findings are also applicable to accelerometry methodology. Therefore, the potential role of physical fitness as a determinant of PAEE is a relevant consideration to all studies that use PAEE estimation for quantifying volume or the intensity distribution of physical activity. Therefore, we encourage the inclusion of both fitness and physical activity assessments in epidemiological studies.

The strengths of the current study include the population-based sample of older men and women, which was reasonably balanced in terms of the sex of the participants including 61.9 % of women, which is comparable to the national proportion of women in this age group (61.3 %) in 2017 (Statistics Finland, 2023). The subsample was somewhat more physically active based on self-report than those who did not volunteer for the device-based monitoring. The subsample also had a faster walking speed (Portegijs et al., 2019). Therefore, we cannot generalise our findings to represent the whole population of older people, which is an important but not unusual limitation when examining this age group.

To conclude, we observed higher volume and intensity of physical activity in a population-based sample of older men compared to women, a difference which was largely determined by sex differences in adiposity and cardiorespiratory fitness. The mediation effect of muscle strength was weaker and significant only when PAEE was assessed using accelerometry only. Our findings stress the importance of keeping fit and maintaining a healthy weight in order to support active living in older adults.

## Author contributions

Laura Karavirta: Conceptualization, Data Curation, Formal Analysis, Funding Acquisition, Investigation, Methodology, Project Administration, Software, Supervision, Visualization, Writing – Original Draft, Writing – Review & Editing

Timo Aittokoski: Data Curation, Formal Analysis, Writing – Review & Editing

Katja Pynnönen: Formal Analysis, Methodology, Writing – Review & Editing

Timo Rantalainen: Formal Analysis, Writing – Review & Editing

Kate Westgate: Software, Writing – Review & Editing

Tomas Gonzales: Software, Writing – Review & Editing

Joona Neuvonen: Data Curation, Writing – Review & Editing

Lotta Palmberg: Data Curation, Writing – Review & Editing

Jukka A. Lipponen: Formal Analysis, Writing – Review & Editing

Katri Turunen: Data Curation, Writing – Review & Editing

Riku Nikander: Data Curation, Writing – Review & Editing

Erja Portegijs: Data Curation, Writing – Review & Editing

Taina Rantanen: Funding Acquisition, Supervision, Writing – Review & Editing

Søren Brage: Methodology, Software, Supervision, Writing – Review & Editing

## Data Availability

Pseudonymised data are available to external collaborators upon reasonable request under the terms of data protection regulation. To request the data please contact Professor Taina Rantanen.

https://doi.org/10.17011/jyx/dataset/83811

## Supporting information

Supplementary file 1 (accelerometry method)

Supplementary file 2 (Tables S2.1 and S2.2)

Supplementary file 3 (Figures S3.1, S3.2, and S3.3)

