## Supplemental file 1 for "Physical determinants of daily physical activity in older men and women"

Linear conversion of acceleration to physical activity energy expenditure

**METHODS**

*Participants.* A convenience sample of older adults (n=12) were invited to perform a treadmill test. Participants were required to provide a written informed consent. Body height and mass were measured using standard calibrated equipment. Fat-free mass and body fat percentage were assessed using multi-frequency bioelectrical impedance measurement (InBody 720, Biospace, Seoul, Korea). Beta-blocker use was self-reported. Preferred walking speed was measured with photocells over 10 meters in a laboratory corridor allowing acceleration and deceleration before and after the start and finish line, respectively.

*Accelerometry.* A tri-axial accelerometer (UKK RM42, 100 Hz, 13-bit ±16g, UKK Terveyspalvelut Oy, Tampere, Finland) was attached on the mid-thigh of the dominant leg. Pre-processing of acceleration to mean amplitude deviation (MAD) was performed similarly as for the free-living AGNES data, except for calibration, which was not possible due to the lack of stationary data in the treadmill protocol. Average MAD minus baseline noise was computed for each walking speed. Baseline noise was defined from the AGNES free-living data (n=409) as the upper bound of 99 % CI of the lowest observed MAD during free-living (0.0033 g).

*Energy expenditure.* VO_2_ and VCO_2_ for walking speeds 1.5 km/h, 3.0 km/h, 4.5 km/h, and 6.0 km/h were measured using indirect calorimetry (Jaeger Oxycon Pro, Viasys Healthcare Gmbh, Hoechberg, Germany). VO_2_ is expressed per kg of body mass and per meter of travelled distance to provide information on the cost of transport on the treadmill. Physical activity energy expenditure (PAEE, in J/kg/min) was calculated using the equation by Weir et al.

4184 * (3.941 * VO_2_ + 1.106 * VCO_2_ – BMR) / body mass,

where VO_2_ and VCO_2_ are 1-minute averages of oxygen uptake and carbon dioxide production from the steady-state portion of each walking speed, and basal metabolic rate (BMR) was estimated using a sex-stratified equation for older adults based on body mass and height: 0.0356 * body mass + 1.76 * height + 0.0448 for women and 0.0478 * body mass + 2.26 * height – 1.07 for men, and transformed to J/min by dividing by 1.44 (Henry, 2005). PAEE at each walking speed was additionally expressed as METs by dividing PAEE by estimated BMR. Standard gross METs were calculated by dividing measured VO_2_ by 3.5 ml/kg/min (Ainsworth et al., 2011).

*Heart rate* was measured using Polar heart rate monitors (Polar Electro Oy, Kempele, Finland) and averaged for the steady-state phase of each walking speed.

*Statistical analysis.* Participant characteristics and treadmill walking data are presented separately for men and women as mean and standard deviation or number and proportion of participants. Men and women were not compared due to the small sample size. Multiple linear regression (using maximum likelihood modelling) with random effects on individual level was used to derive conversion equation for MAD to PAEE (Brage et al., 2007) using Stata 16.0 (StataCorp LLC, Texas, U.S.A.).

**RESULTS**

Participant characteristics are reported in Table S1.1, and data from the treadmill test in Table S1.2.

**Table S1.1**. Participant characteristics.

|  | All (N=12) | Men (n=6) | Women (n=6) |
| --- | --- | --- | --- |
| Age (yr) | 71.2 (5.0) | 72.3 (5.1) | 70.0 (5.0) |
| Height (m) | 1.67 (0.09) | 1.74 (0.03) | 1.59 (0.06) |
| Body mass (kg) | 74.6 (15.3) | 78.7 (11.2) | 70.6 (18.6) |
| BMI (kg/m^2^) | 26.8 (4.8) | 25.9 (3.2) | 27.8 (6.2) |
| FFM (kg) | 50.4 (9.5) | 58.2 (3.4) | 42.5 (6.3) |
| Body fat (%) | 31.7 (9.9) | 25.4 (6.1) | 38.0 (9.0) |
| Walk speed (km/h) | 5.0 (0.4) | 5.0 (0.2) | 5.0 (0.5) |
| BMR (J/kg/min) | 56.5 (5.5) | 58.9 (3.0) | 54.1 (6.7) |
| Betablocker (n/%) | 3 (25.0) | 2 (33.3) | 1 (16.7) |

BMI, body mass index; FFM, fat-free mass; BMR, estimated basal metabolic rate.

**Table S1.2.** Treadmill walking.

| Speed (km/h) | VO_2_ (ml/kg/min) | VO_2_ (ml/kg/m) | PAEE (J/kg/min) | PAEE (stdMET) | PAEE (bmrMET) | HR (bpm) | MAD (g) |
| --- | --- | --- | --- | --- | --- | --- | --- |
| 1.5 | 8.0 (1.3) | 0.32 (0.05) | 108.0 (28.7) | 1.3 (0.4) | 1.9 (0.6) | 79.5 (13.3) | 0.076 (0.046) |
| 3.0 | 10.8 (2.2) | 0.22 (0.04) | 164.5 (47.2) | 2.1 (0.6) | 3.0 (1.0) | 88.1 (11.5) | 0.224 (0.068) |
| 4.5 | 12.7 (2.5) | 0.17 (0.03) | 203.0 (52.3) | 2.6 (0.7) | 3.6 (1.0) | 94.9 (12.5) | 0.423 (0.074) |
| 6.0 | 17.2 (2.4) | 0.17 (0.02) | 299.6 (52.5) | 3.9 (0.7) | 5.4 (1.2) | 112.7 (16.1) | 0.671 (0.124) |

PAEE, physical activity energy expenditure; stdMET, standard MET (3.5 ml/kg/min) above rest; bmrMET, multiples of individually estimated BMR using Henry equations, above rest; HR, heart rate; MAD, mean amplitude deviation in g (Earth’s gravity).

MAD was significantly related to PAEE (Z=14.16, p<0.001). The estimated slope (β_1_) was 295.3 and the intercept (β_0_) was 90.8. The model accounted for 72.0 % of the variance.

Therefore, the equation to predict individual PAEE from MAD was,

PAEE (J/kg/min) = 295.3 * MAD + 90.8

The individual data points and the resulting linear equation are presented in Figure S1.1. The standard 2 net MET cut-point for moderate (3 gross METs or PAEE 142 J/kg/min) and 5 net MET cut-point for vigorous (6 gross METs or PAEE 356 J/kg/min) physical activity using the equation were 0.175 g and 0.899 g.

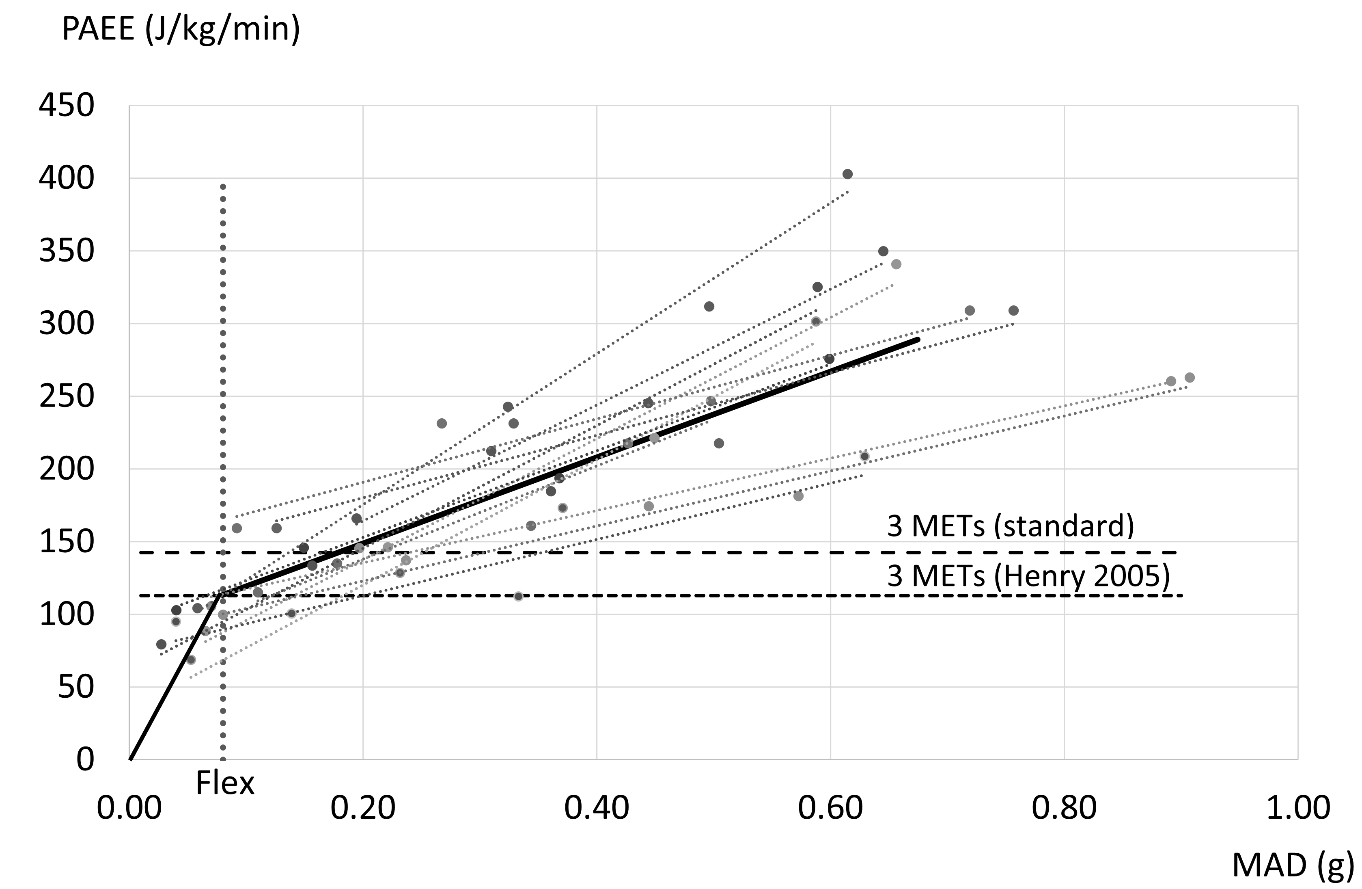

**Figure S1.1**. Linear association between mean amplitude deviation (MAD) and physical activity energy expenditure (PAEE) at the individual (dotted lines) and group level (solid line).
