## Supplemental file 2 for "Physical determinants of daily physical activity in older men and women"

**Table S2.1.** Results of path modelling testing components of physical fitness as mediators in the relationships between sex and daily physical activity energy expenditure (PAEE), moderate-to-vigorous physical activity (MVPA), and light physical activity (LPA) (n=409).

|  | **Associations between the model variables** | | | | |  | **Indirect effects** | | | | |
| --- | --- | --- | --- | --- | --- | --- | --- | --- | --- | --- | --- |
|  | **B** | **SE** | **β** | **95 % CI** | **p** |  | **B** | **SE** | **β** | **95 % CI** | **p** |
| **Outcome: PAEE** |  |  |  |  |  |  |  |  |  |  |  |
| Sex→PAEE | -0.13 | 1.13 | -0.01 | -0.12; 0.11 | .911 |  |  |  |  |  |  |
| Sex→Body fat (%) | -11.75 | 0.69 | -0.63 | -0.69; -0.58 | <.001 |  |  |  |  |  |  |
| Body fat (%)→PAEE | -0.33 | 0.07 | -0.31 | -0.43; -0.19 | <.001 |  | 3.91 | 0.82 | 0.20 | 0.12;0.27 | <.001 |
| Sex→Walk speed | 0.28 | 0.07 | 0.17 | 0.09; 0.26 | <.001 |  |  |  |  |  |  |
| Walk speed→PAEE | 3.29 | 0.63 | 0.27 | 0.17; 0.36 | <.001 |  | 0.92 | 0.30 | 0.05 | 0.02;0.08 | .001 |
| Sex→ Muscle strength | 1.44 | 0.13 | 0.47 | 0.40; 0.54 | <.001 |  |  |  |  |  |  |
| Muscle strength→PAEE | 0.40 | 0.38 | 0.06 | -0.05; 0.17 | .279 |  | 0.58 | 0.52 | 0.03 | -0.02;0.08 | .269 |
| Body fat (%) with Walk speed | -1.77 | 0.25 | -0.37 | -0.45; -0.28 | <.001 |  |  |  |  |  |  |
| Body fat (%) with Muscle strength | -3.69 | 0.48 | -0.43 | -0.51; -0.35 | <.001 |  |  |  |  |  |  |
| Walk speed with Muscle strength | 0.40 | 0.04 | 0.45 | 0.37; 0.53 | <.001 |  |  |  |  |  |  |
| Chronic conditions→PAEE | -0.73 | 0.22 | -0.14 | -0.22; -0.06 | .001 |  |  |  |  |  |  |
| Chronic conditions→Body fat (%) | 0.74 | 0.20 | 0.15 | 0.07; 0.23 | .001 |  |  |  |  |  |  |
| Age→Walk speed | -0.05 | 0.01 | -0.20 | -0.31; -0.10 | <.001 |  |  |  |  |  |  |
| Chronic conditions→Walk speed | -0.13 | 0.02 | -0.30 | -0.38; -0.21 | <.001 |  |  |  |  |  |  |
| Duration of education→Walk speed | 0.02 | 0.01 | 0.09 | 0.02; 0.17 | .010 |  |  |  |  |  |  |
| Age→Muscle strength | -0.07 | 0.02 | -0.15 | -0.31; -0.10 | <.001 |  |  |  |  |  |  |
| Chronic conditions→Muscle strength | -0.13 | 0.04 | -0.16 | -0.24; -0.07 | .001 |  |  |  |  |  |  |
| Age with Duration of education | -1.86 | 0.66 | -0.13 | -0.22; -0.04 | .003 |  |  |  |  |  |  |
| Age with Chronic conditions | 1.10 | 0.30 | 0.18 | 0.09; 0.27 | <.001 |  |  |  |  |  |  |
| **Outcome: MVPA** |  |  |  |  |  |  |  |  |  |  |  |
| Sex→MVPA | 0.34 | 0.76 | 0.03 | -0.09; 0.12 | .656 |  |  |  |  |  |  |
| Sex→Body fat (%) | -11.76 | 0.69 | -0.63 | -0.69; -0.58 | <.001 |  |  |  |  |  |  |
| Body fat (%)→MVPA | -0.13 | 0.04 | -0.17 | -0.28; -0.07 | .001 |  | 1.49 | 0.55 | 0.11 | 0.04;0.18 | .002 |
| Sex→Walk speed | 0.28 | 0.07 | 0.17 | 0.09; 0.26 | <.001 |  |  |  |  |  |  |
| Walk speed→MVPA | 2.36 | 0.43 | 0.28 | 0.19; 0.37 | <.001 |  | 0.66 | 0.21 | 0.05 | 0.02;0.08 | .001 |
| Sex→Muscle strength | 1.44 | 0.13 | 0.47 | 0.40; 0.54 | <.001 |  |  |  |  |  |  |
| Muscle strength→MVPA | 0.45 | 0.25 | 0.10 | -0.01; 0.21 | .074 |  | 0.65 | 0.35 | 0.05 | -0.00;0.10 | .068 |
| Body fat (%) with Walk speed | -1.76 | 0.25 | -0.37 | -0.45; -0.28 | <.001 |  |  |  |  |  |  |
| **Outcome: MVPA** |  |  |  |  |  |  |  |  |  |  |  |
| Body fat (%) with Muscle strength | -3.69 | 0.48 | -0.43 | -0.51; -0.35 | <.001 |  |  |  |  |  |  |
| Walk speed with Muscle strength | 0.40 | 0.04 | 0.45 | 0.37; 0.53 | <.001 |  |  |  |  |  |  |
| Chronic conditions→MVPA | -0.46 | 0.14 | -0.13 | -0.21; -0.05 | .001 |  |  |  |  |  |  |
| Chronic conditions→Body fat (%) | 0.74 | 0.20 | 0.15 | 0.07; 0.23 | <.001 |  |  |  |  |  |  |
| Age→Walk speed | -0.05 | 0.01 | -0.20 | -0.30; -0.10 | <.001 |  |  |  |  |  |  |
| Chronic conditions→Walk speed | -0.13 | 0.02 | -0.30 | -0.38; -0.21 | <.001 |  |  |  |  |  |  |
| Duration of education→Walk speed | 0.02 | 0.01 | 0.09 | 0.02; 0.16 | .010 |  |  |  |  |  |  |
| Age→Muscle strength | -0.07 | 0.02 | -0.15 | -0.23; -0.08 | <.001 |  |  |  |  |  |  |
| Chronic conditions→Muscle strength | -0.13 | 0.02 | -0.16 | -0.24: -0.07 | .001 |  |  |  |  |  |  |
| Age with Duration of education | -1.86 | 0.66 | -0.13 | -0.22: -0.04 | .003 |  |  |  |  |  |  |
| Age with Chronic conditions | 1.10 | 0.30 | 0.18 | 0.09; 0.27 | <.001 |  |  |  |  |  |  |
| **Outcome: LPA** |  |  |  |  |  |  |  |  |  |  |  |
| Sex→LPA | -0.34 | 0.66 | -0.03 | -0.16; 0.09 | .605 |  |  |  |  |  |  |
| Sex→ Body fat (%) | -11.76 | 0.69 | -0.63 | -0.69; -0.57 | <.001 |  |  |  |  |  |  |
| Body fat (%)→LPA | -0.22 | 0.04 | -0.38 | -0.52; -0.25 | <.001 |  | 2.66 | 0.50 | 0.24 | 0.15;0.34 | <.001 |
| Sex→ Walk speed | 0.28 | 0.07 | 0.17 | 0.09; 0.26 | <.001 |  |  |  |  |  |  |
| Walk speed→ LPA | 0.99 | 0.35 | 0.15 | 0.05; 0.26 | .005 |  | 0.28 | 0.12 | 0.03 | 0.00;0.05 | .019 |
| Sex→ Muscle strength | 1.44 | 0.13 | 0.47 | 0.40; 0.54 | <.001 |  |  |  |  |  |  |
| Muscle strength→LPA | -0.13 | 0.21 | -0.04 | -0.16; 0.08 | .555 |  | -0.18 | 0.31 | -0.02 | -0.08;0.04 | .560 |
| Body fat (%) with Walk speed | -1.76 | 0.25 | -0.37 | -0.45; -0.28 | <.001 |  |  |  |  |  |  |
| Body fat (%) with Muscle strength | -3.68 | 0.48 | -0.43 | -0.51; -0.35 | <.001 |  |  |  |  |  |  |
| Walk speed with Muscle strength | 0.39 | 0.04 | 0.45 | 0.37; 0.53 | <.001 |  |  |  |  |  |  |
| Chronic conditions→LPA | -0.28 | 0.13 | -0.10 | -0.19; -0.01 | .032 |  |  |  |  |  |  |
| Chronic conditions→Body fat (%) | 0.74 | 0.20 | 0.15 | 0.07: 0.23 | <.001 |  |  |  |  |  |  |
| Age→Walk speed | -0.05 | 0.01 | -0.20 | -0.31; -0.10 | <.001 |  |  |  |  |  |  |
| Chronic conditions→Walk speed | -0.13 | 0.02 | -0.30 | -0.38; -0.21 | <.001 |  |  |  |  |  |  |
| Duration of education→Walk speed | 0.02 | 0.01 | 0.09 | 0.02; 0.16 | .010 |  |  |  |  |  |  |
| Age→Muscle strength | -0.07 | 0.02 | -0.16 | -0.23; -0.08 | <.001 |  |  |  |  |  |  |
| Chronic conditions→Muscle strength | -0.13 | 0.04 | -0.15 | -0.24; -0.07 | .001 |  |  |  |  |  |  |
| Age with Duration of education | -1.86 | 0.66 | -0.13 | -0.22: -0.04 | .003 |  |  |  |  |  |  |
| Age with Chronic conditions | 1.10 | 0.30 | 0.18 | 0.09; 0.27 | <.001 |  |  |  |  |  |  |

**Table S2.2.** Results of sensitivity analysis of path modelling testing components of physical fitness as mediators in the relationship between sex and physical activity energy expenditure measured using accelerometry (PAEE_ACC) (n=409).

|  | **Associations between the model variables** | | | | |  | **Indirect effects** | | | | |
| --- | --- | --- | --- | --- | --- | --- | --- | --- | --- | --- | --- |
|  | **B** | **SE** | **β** | **95 % CI** | **p** |  | **B** | **SE** | **β** | **95 % CI** | **p** |
| Sex→ PAEE_ACC | -0.91 | 0.99 | -0.06 | -0.18; 0.06 | .349 |  |  |  |  |  |  |
| Sex→ Body fat (%) | -11.77 | 0.69 | -0.64 | -0.69; -0.58 | <.001 |  |  |  |  |  |  |
| Body fat (%)→PAEE_ACC | -0.22 | 0.06 | -0.26 | -0.38; -0.13 | <.001 |  | 2.60 | 0.73 | 0.16 | 0.08;0.25 | <.001 |
| Sex→ Walk speed | 0.28 | 0.07 | 0.17 | 0.09; 0.26 | <.001 |  |  |  |  |  |  |
| Walk speed→ PAEE_ACC | 2.56 | 0.50 | 0.26 | 0.16; 0.36 | <.001 |  | 0.72 | 0.24 | 0.05 | 0.02;0.07 | .002 |
| Sex→ Muscle strength | 1.44 | 0.13 | 0.47 | 0.40; 0.54 | <.001 |  |  |  |  |  |  |
| Muscle strength→ PAEE_ACC | 0.57 | 0.30 | 0.11 | 0.00; 0.22 | .047 |  | 0.82 | 0.41 | 0.05 | 0.00;0.10 | .042 |
| Body fat (%) with Walk speed | -1.77 | 0.25 | -0.37 | -0.45; -0.29 | <.001 |  |  |  |  |  |  |
| Body fat (%) with Muscle strength | -3.69 | 0.48 | -0.43 | -0.51: -0.35 | <.001 |  |  |  |  |  |  |
| Walk speed with Muscle strength | 0.39 | 0.04 | 0.45 | 0.37; 0.53 | <.001 |  |  |  |  |  |  |
| Chronic conditions→PAEE_ACC | -0.65 | 0.19 | -0.15 | -0.24; -0.07 | .001 |  |  |  |  |  |  |
| Chronic conditions→Body fat (%) | 0.75 | 0.20 | 0.15 | 0.07; 0.23 | <.001 |  |  |  |  |  |  |
| Age→Walk speed | -0.05 | 0.01 | -0.20 | -0.31; -0.10 | <.001 |  |  |  |  |  |  |
| Chronic conditions→Walk speed | -0.13 | 0.02 | -0.30 | -0.38; -0.21 | <.001 |  |  |  |  |  |  |
| Duration of education→Walk speed | 0.02 | 0.01 | 0.09 | 0.02; 0.16 | .010 |  |  |  |  |  |  |
| Age→Muscle strength | -0.07 | 0.02 | -0.15 | -0.23; -0.08 | <.001 |  |  |  |  |  |  |
| Chronic conditions→Muscle strength | -0.13 | 0.02 | -0.16 | -0.24; -0.07 | .001 |  |  |  |  |  |  |
| Age with Duration of education | -1.86 | 0.66 | -0.13 | -0.22: -0.04 | .003 |  |  |  |  |  |  |
| Age with Chronic conditions | 1.10 | 0.30 | 0.18 | 0.09; 0.27 | <.001 |  |  |  |  |  |  |
