## Supplemental file 3 for "Physical determinants of daily physical activity in older men and women"

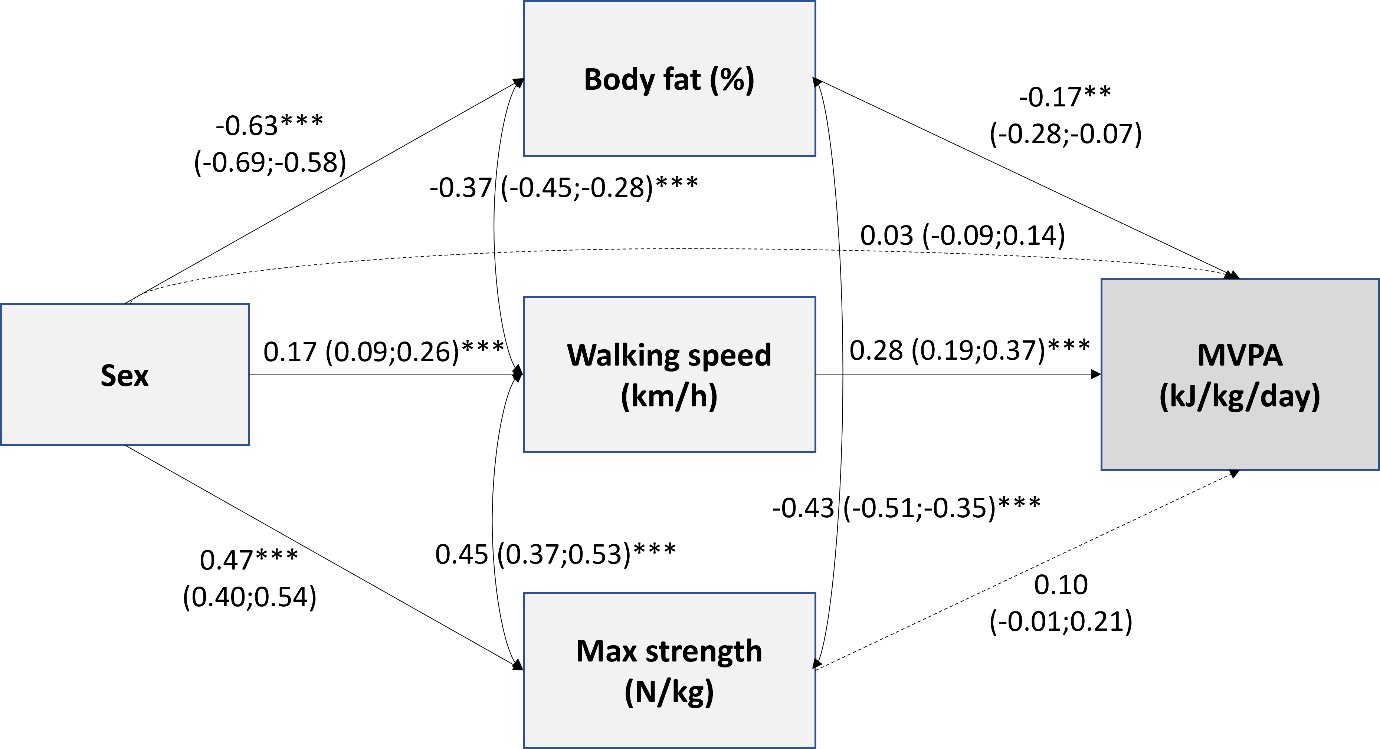

**Fig.S3.1.** Standardized beta coefficients (with 95% confidence intervals) from path model testing factors describing physical fitness (body fat percentage, walking speed, and muscle strength) as mediators in the relationships between sex and moderate-to-vigorous physical activity (MVPA) energy expenditure (*p≤0.05, **p≤0.01, ***p≤0.001).

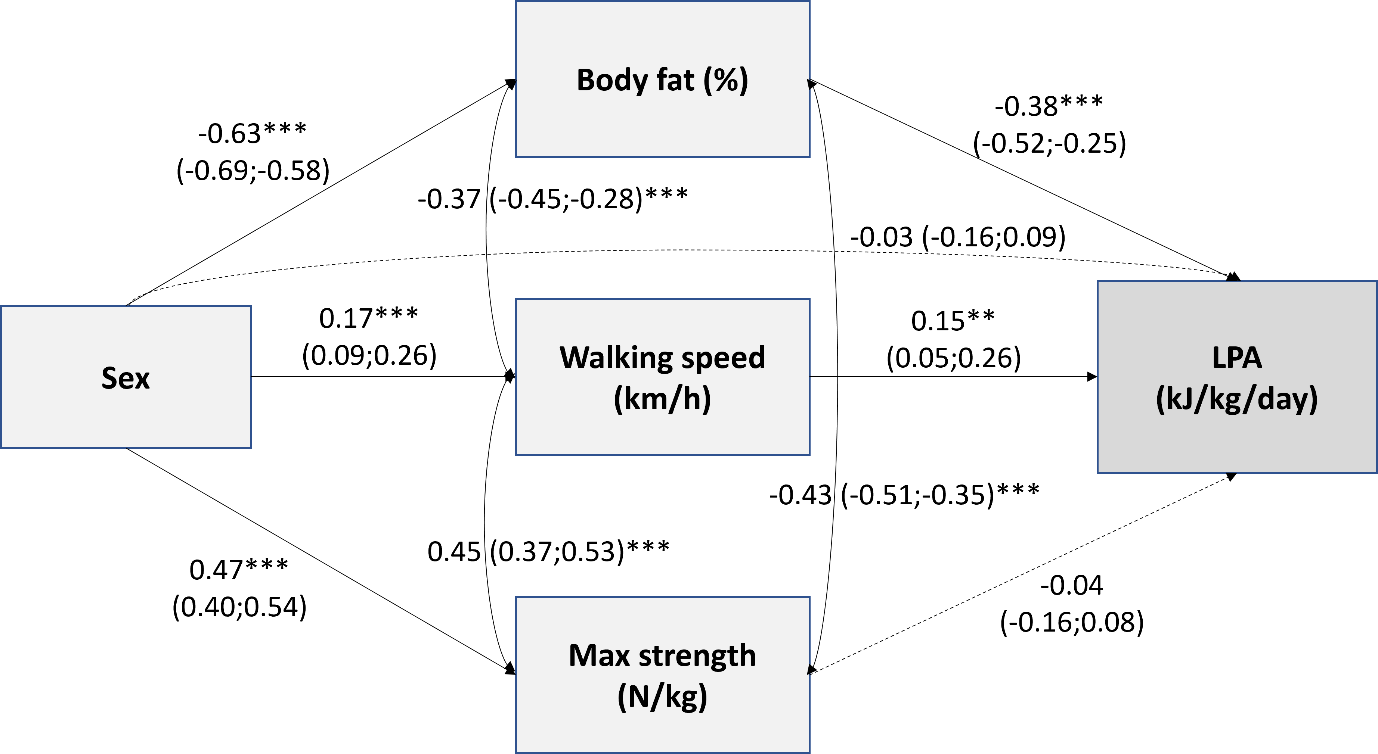

**Fig.S3.2.** Standardized beta coefficients (with 95% confidence intervals) from structural equation model testing factors describing physical fitness (body fat percentage, walking speed, and muscle strength) as mediators in the relationships between sex and light-intensity physical activity (LPA) energy expenditure (*p≤0.05, **p≤0.01, ***p≤0.001).

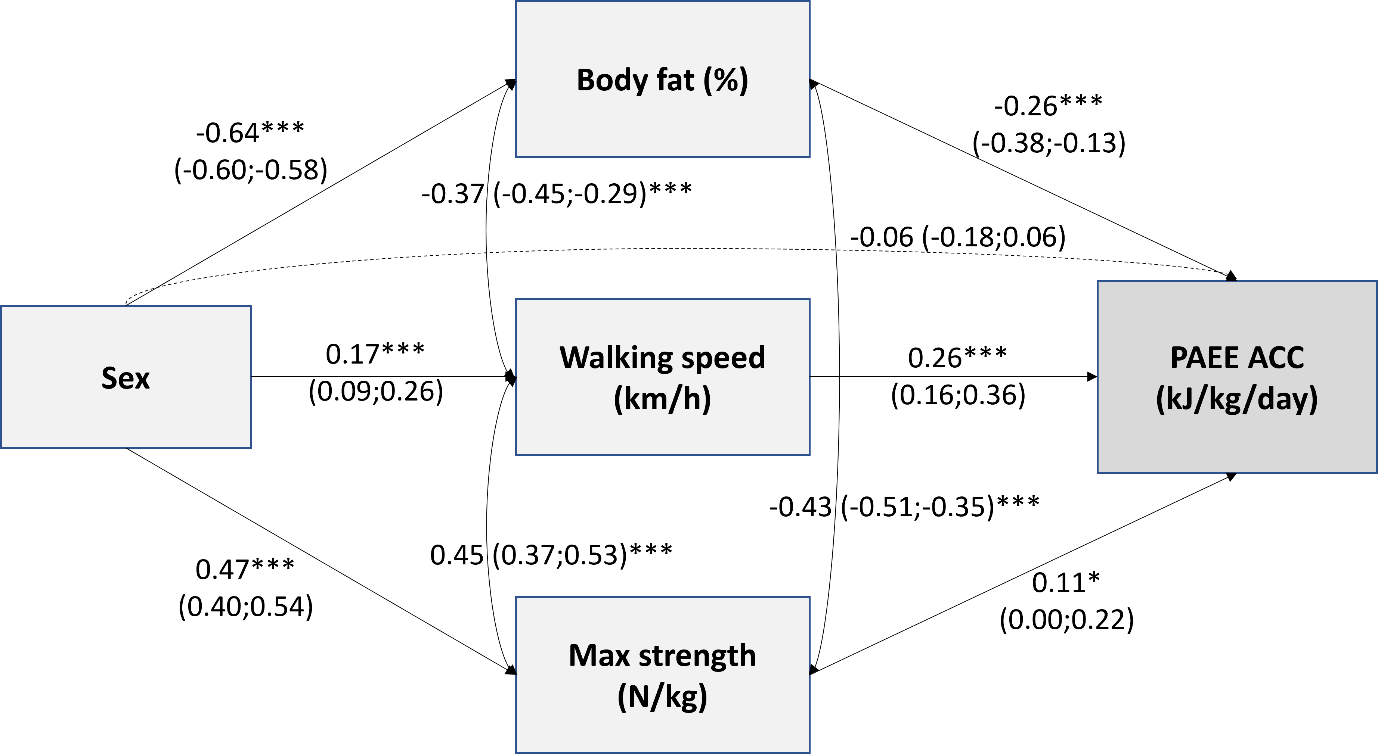
**Fig.S3.3.** Standardized beta coefficients (with 95% confidence intervals) from structural equation model testing factors describing physical fitness (body fat percentage, walking speed, and muscle strength) as mediators in the relationships between sex and physical activity energy expenditure (PAEE) assessed based on accelerometry (ACC) only (*p≤0.05, **p≤0.01, ***p≤0.001).
